# Impact of stopwatch use on propofol administration in upper gastrointestinal endoscopy: A propensity score matching analysis

**DOI:** 10.1101/2023.07.21.23292749

**Authors:** Yong bae Kim

## Abstract

**Background and Aims:** Sedation endoscopy using propofol significantly reduces procedure-related discomforts and increases patient compliance. However, propofol has a very narrow therapeutic window, and the dose required for each individual is very diverse; hence, it must be used with extreme caution. Maintaining an appropriate dosing interval is also important to administer the appropriate dose. This study aimed to investigate the change in propofol dose before and after stopwatch installation in the endoscopy unit and to assess the effect of administration intervals on propofol usage patterns.

**Methods:** This retrospective study included people who underwent sedation endoscopy without biopsies between January 1 and October 31 of 2019. The participants were divided into before (n = 526) and after (n = 845) groups. In the after group, drug interval was rigorously maintained using the stopwatch. Changes in propofol dose with respect to sex, age, weight, height, and endoscopic procedure time were statistically analyzed. Group difference between each variable was reduced by propensity score matching adjustment, and statistical analysis was conducted using the R program.

**Results:** No adverse event occurred. The propofol dosage per body weight decreased from 1.38 mg/kg (before) to 1.27 mg/kg (after) in females (*p* < 0.0001), while it increased from 1.23 mg/kg to 1.28 mg/kg in males (*p* = 0.016). These changes were found to be significantly different between sexes before the installation of the stopwatch (*p* < 0.0001). However, after the installation of the stopwatch, there was no significant difference in propofol dosage between males and females (1.28 mg/kg vs. 1.27 mg/kg; *p* = 0.9752).

**Conclusions:** The use of a stopwatch in sedative upper gastrointestinal endoscopy reduced the propofol requirement in females but not in males.

## Introduction

Upper gastrointestinal endoscopy is important for the early detection of gastrointestinal diseases, but it may cause considerable discomfort, such as nausea and pain, making patients reluctant to undergo such procedure. However, sedation endoscopy significantly reduces these drawbacks and improve patient compliance [1–3].

Propofol is a beneficial choice for screening with upper gastrointestinal endoscopy due to its fast-acting and short duration properties. This is particularly advantageous for the brief nature of the procedure. Furthermore, propofol has fewer side effects, including a reduced risk of residual “hangover” effects, compared to opioids or short-acting benzodiazepines [4–9].

However, it has a minimal analgesic effect, has a lower amnesic effect than midazolam, and causes vascular pain when administered [10]. Furthermore, propofol overdose may result in respiratory or cardiac dysfunction, and it has no antidote, making this drug potentially dangerous [11,12]. Due to the narrow therapeutic window and significant inter-individual variability in required doses, utmost caution is necessary to ensure safe and appropriate use of propofol [13,14]. Thus, in many European countries, the anesthesiologist should control the patient’s level of sedation during most of the endoscopy [15,16].

In other countries, however, the endoscopist alone safely adjusts the level of sedation with the help of a specially educated nurse [17,18]. While endoscopists can safely manage a patient’s state of consciousness with propofol, drug administration errors remain possible. For example, if propofol is administered several times at a very short interval, it can be excessively delivered [19]. Therefore, keeping an appropriate dosing interval while checking patient’s response is important for safe dose administration. In situations where unpredictable events occur, such as a paradoxical reaction to the sedative, the endoscopist might experience feelings of anxiety or nervousness. This psychological burden can potentially disrupt the endoscopist’s perception of time, leading to possible deviations in the drug administration interval. [20,21].

In most cases, relying on a standard clock is adequate for monitoring the passage of time and maintaining appropriate intervals between drug administrations. However, if the endoscopist becomes distracted by an unforeseen event, such as the aforementioned paradoxical reaction, accurately perceiving the passage of time with a regular clock may be challenging. Therefore, it would be reasonable to introduce a more intuitive method to track the dosing interval in the endoscopy room, such as a wall-mounted stopwatch with clear visibility.

This study aimed to investigate the change in propofol dose before and after stopwatch installation in the endoscopy unit and to assess the effect of administration intervals on propofol usage patterns.

## Materials and Methods

In this retrospective study, patients over the age of 20 who underwent sedation endoscopy at the Health Care Center of Namcheon Hospital in Gyeonggi-do, Korea in 2019 were included as participants. Participants were categorized into two groups, namely the “before” group (January 1 to May 31) and the “after” group (July 1 to October 31), based on the introduction of the wall-mounted digital stopwatch in the endoscopy unit (Fig. 1), with June acting as the transitional period.

**Figure 1.**
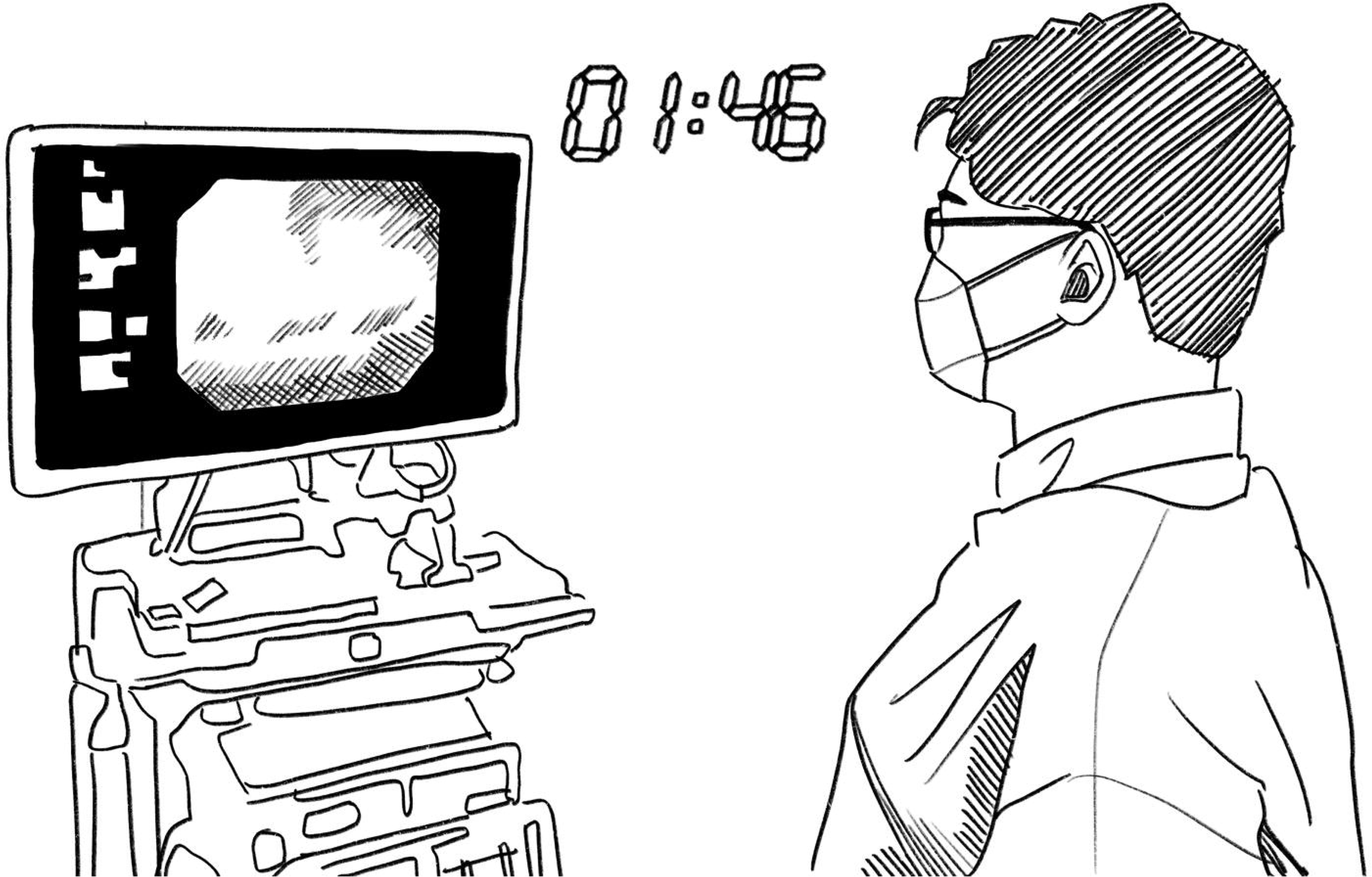
The location of the stopwatch installed in the endoscopy unit.

Data on propofol dosage, as well as information on sex, age, weight, height, and endoscopic procedure time, were collected from the medical records. The endoscopic procedure time was defined as the duration between the imaging time of the first image (esophagogastric junction image) and the time of the last image captured during the procedure. Those who underwent biopsy, a factor that could interfere with analysis because of prolonged endoscopic procedures, were excluded.

As previous studies have identified significant differences in the dosage of propofol administered based on sex, a gender-specific analysis was performed in both groups to account for potential disparities in propofol requirement between males and females [22–24]. To mitigate the heterogeneity among the patient groups, the MatchIt library in R software was used to perform propensity score matching for adjustment between the two groups (1:1 nearest-neighbor method). (Fig. 2)

**Figure 2.**
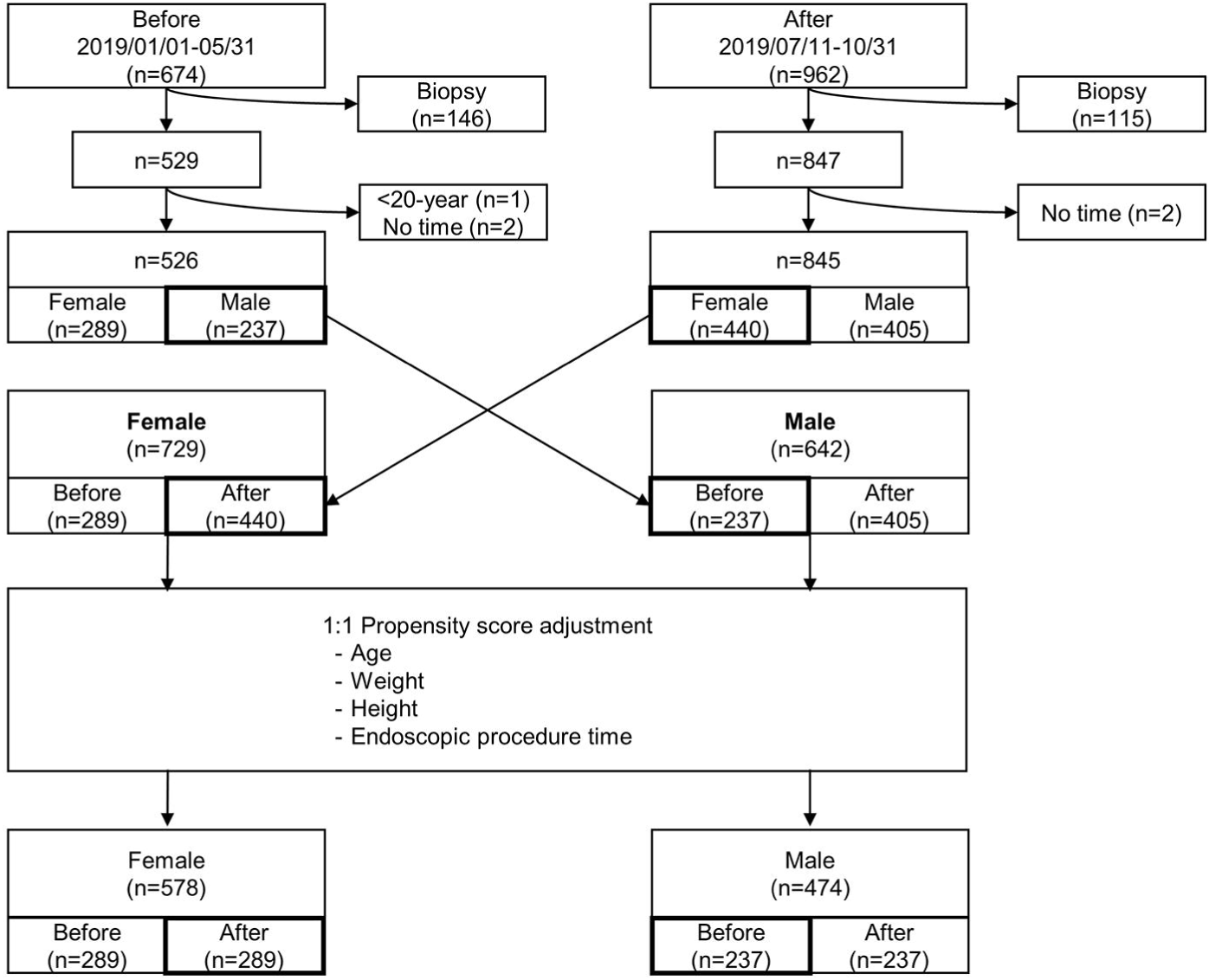
Study data collection and 1:1 propensity score matching adjustment.

After obtaining ethical approval from the Public Institutional Review Board designated by South Korea Ministry of Health and Welfare (P01-202102-11-001), an anonymized patient case database was established on February 17, 2021, to support subsequent analysis, and access to the data was made available. The author performed all endoscopy procedures, recording, data collection, and analysis. During the data collection phase, the author had access to information that could identify individual participants. However, after the establishment of the anonymized patient database, the author no longer had access to any information that could identify individual participants.

All examinees received lidocaine (Xylocaine) pump spray (10mg/pump) 1–2 times in the throat for local anesthesia before the start of the endoscopy. No other premedication was administered. During the endoscopic procedure, patients’ oxygen saturation was monitored, and they were administered oxygen at a rate of 3 liters per minute through a nasal cannula.

The initial dose of propofol (Anepol injection [120 mg]; Hana Pharm Co., Ltd.) ranged from 0.5 to 1.2 mg/kg, considering the age of the participant. If additional doses were required, 10–20 mg dose was repeatedly administered until the desired level of sedation was reached, but the injected dose was not more than 20 mg at a time. In the “before” group, the endoscopist was allowed to decide the timing of additional propofol administration at their discretion. However, in the “after” group, the procedure was more systematic: a stopwatch was initiated immediately after the initial propofol administration. After a 30-second period (reflecting the arm-brain circulation time), the endoscopist would then decide whether to proceed with the endoscopy or administer an additional dose of propofol. If more propofol was needed, the interval between injections should be at least 20–30 seconds.

### Statistical analysis

The chi-square test was used for the demographic analysis of each group. For the group with a parametric distribution, the mean values were compared by student’s t-test, and for the nonparametric data, the median values were compared by Mann–Whitney *U* test. A two-sided *p*-value less than 0.05 were considered statistically significant. All statistical data were analyzed using the R software version 4.0.0 (R Foundation for Statistical Computing).

## Results and discussion

In this study, no patients were encountered who could not complete the test or required resuscitation due to side effects. In the “before” group, a total of 526 individuals underwent sedation gastroscopy, while in the “after” group, 845 individuals underwent the same procedure. The height and endoscopic procedure time for each group was significantly different (Table 1).

**TABLE 1.**
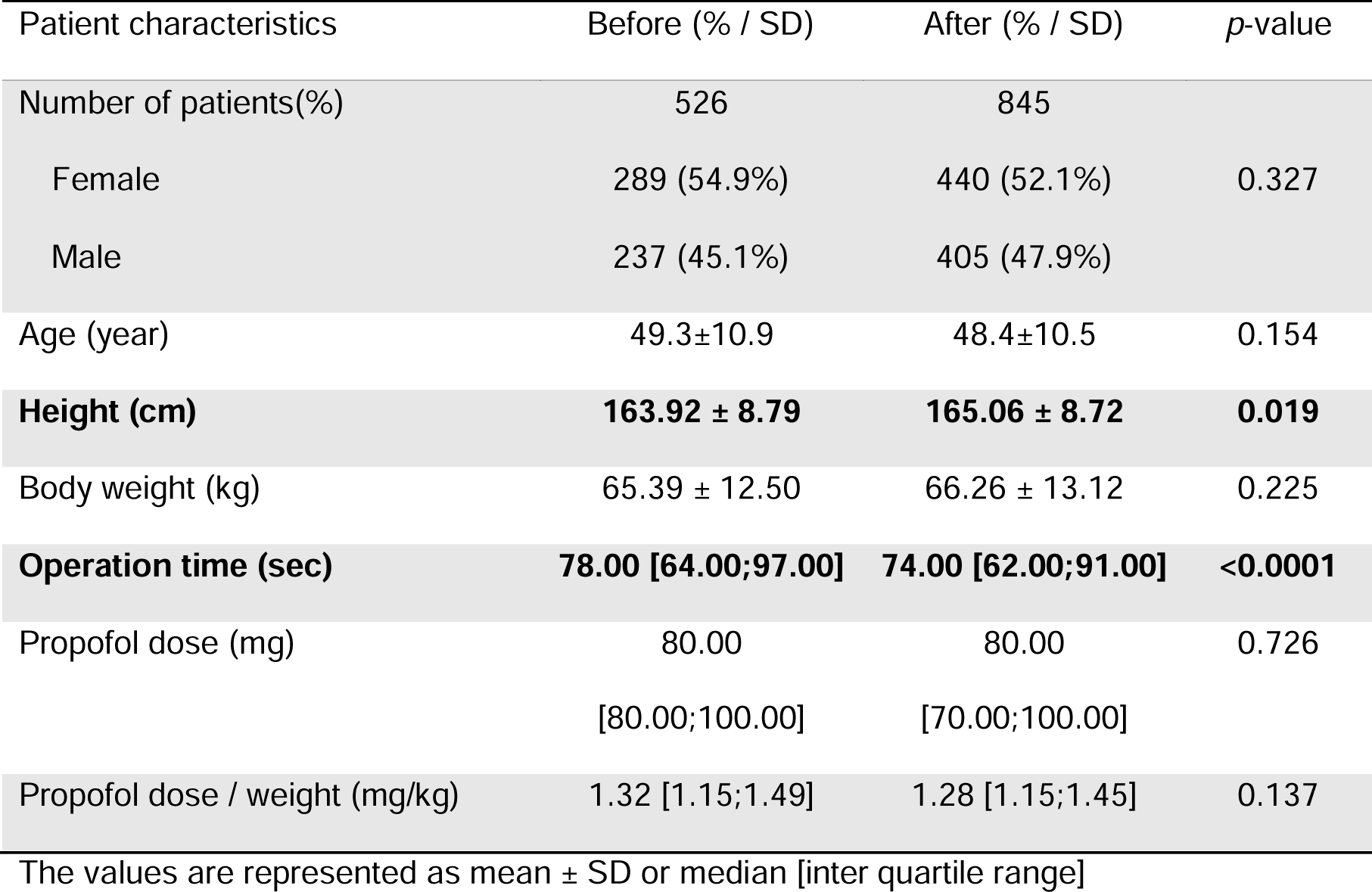
Baseline characteristics.

| Patient characteristics | Before (% / SD) | After (% / SD) | p-value |
| --- | --- | --- | --- |
| Number of patients(%) | 526 | 845 |  |
| Female | 289 (54.9%) | 440 (52.1%) | 0.327 |
| Male | 237 (45.1%) | 405 (47.9%) |  |
| Age (year) | 49.3±10.9 | 48.4±10.5 | 0.154 |
| **Height (cm)** | **163.92 ± 8.79** | **165.06 ± 8.72** | **0.019** |
| Body weight (kg) | 65.39 ± 12.50 | 66.26 ± 13.12 | 0.225 |
| **Operation time (sec)** | **78.00 [64.00;97.00]** | **74.00 [62.00;91.00]** | **<0.0001** |
| Propofol dose (mg) | 80.00 | 80.00 | 0.726 |
|  | [80.00;100.00] | [70.00;100.00] |  |
| Propofol dose / weight (mg/kg) | 1.32 [1.15;1.49] | 1.28 [1.15;1.45] | 0.137 |

The values are represented as mean ± SD or median [inter quartile range]

Before propensity score matching adjustment, the procedure time was significantly different between the “before” and “after” groups in both sexes (Table 2). To resolve this inequality, this study used propensity score matching adjustment with MatchIt library for R software. The propensity scores of the factors such as age, weight, height, and endoscopic procedure time were calculated and matched one by one using the nearest method in males and females separately. After matching, the significant difference between each variable in each group disappeared (Table 3).

**TABLE 2.**
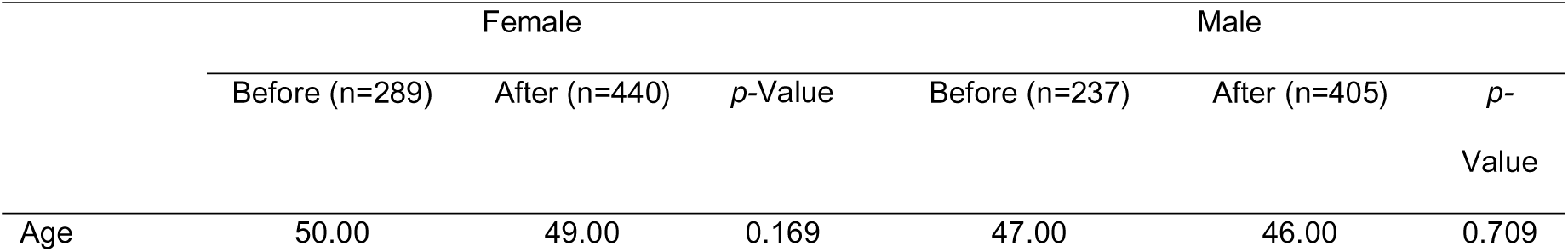
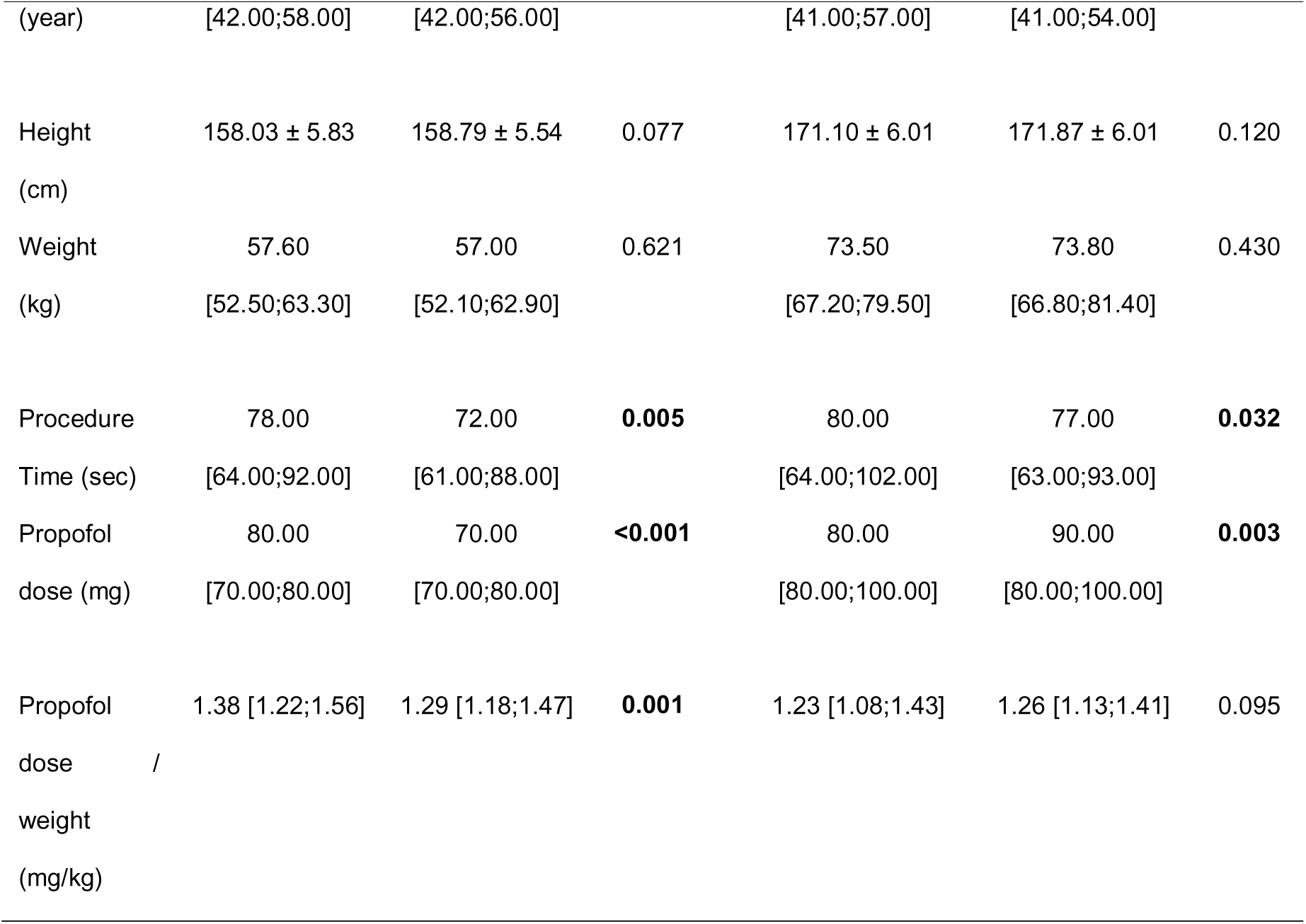
Before propensity score matching.

|  | Female |  |  | Male |  |  |
| --- | --- | --- | --- | --- | --- | --- |
|  | Before (n=289) | After (n=440) | p-Value | Before (n=237) | After (n=405) | p-Value |
| Age | 50.00 | 49.00 | 0.169 | 47.00 | 46.00 | 0.709 |
| (year) | [42.00;58.00] | [42.00;56.00] |  | [41.00;57.00] | [41.00;54.00] |  |
| Height<br>(cm) | 158.03 ± 5.83 | 158.79 ± 5.54 | 0.077 | 171.10 ± 6.01 | 171.87 ± 6.01 | 0.120 |
| Weight<br>(kg) | 57.60<br>[52.50;63.30] | 57.00<br>[52.10;62.90] | 0.621 | 73.50<br>[67.20;79.50] | 73.80<br>[66.80;81.40] | 0.430 |
| Procedure<br>Time (sec) | 78.00<br>[64.00;92.00] | 72.00<br>[61.00;88.00] | **0.005** | 80.00<br>[64.00;102.00] | 77.00<br>[63.00;93.00] | **0.032** |
| Propofol<br>dose (mg) | 80.00<br>[70.00;80.00] | 70.00<br>[70.00;80.00] | **<0.001** | 80.00<br>[80.00;100.00] | 90.00<br>[80.00;100.00] | **0.003** |
| Propofol<br>dose /<br>weight<br>(mg/kg) | 1.38 [1.22;1.56] | 1.29 [1.18;1.47] | **0.001** | 1.23 [1.08;1.43] | 1.26 [1.13;1.41] | 0.095 |

**TABLE 3.** After propensity score matching.

|  | Female |  |  | Male |  |  |
| --- | --- | --- | --- | --- | --- | --- |
|  | Before (n=289) | After (n=289) | p-Value | Before (n=237) | After (n=237) | p-Value |
| Age | 50.00<br>[42.00;58.00] | 50.00<br>[43.00;58.00] | 0.918 | 47.00<br>[41.00;57.00] | 48.00<br>[42.00;56.00] | 0.815 |
| Height<br>(cm) | 158.03 ± 5.83 | 157.98 ± 5.53 | 0.908 | 171.10 ± 6.01 | 171.06 ± 6.01 | 0.929 |
| Weight<br>(kg) | 57.60<br>[52.50;63.30] | 57.20<br>[52.40;63.80] | 0.753 | 73.50<br>[67.20;79.50] | 72.90<br>[65.50;79.00] | 0.441 |
| Procedure Time<br>(sec) | 78.00<br>[64.00;92.00] | 75.00<br>[63.00;92.00] | 0.200 | 80.00<br>[64.00;102.00] | 81.00<br>[65.00;99.00] | 0.909 |
| Propofol dose | 80.00 | 70.00 | **<0.001** | 80.00 | 90.00 | **0.010** |
| (mg) | [70.00;80.00] | [70.00;80.00] |  | [80.00;100.00] | [80.00;100.00] |  |
| Propofol dose / | 1.38 [1.22;1.56] | 1.27 | **<0.001** | 1.23 [1.08;1.43] | 1.28 [1.14;1.45] | **0.016** |
| weight (mg/kg) |  | [1.16;1.45] |  |  |  |  |

Based on these matched data, the PW in each group was compared between males and females, and it was significantly decreased in females but increased in males. PW between sexes was significantly different before stopwatch installation (*p* < 0.001) but showed no statistically significant difference after stopwatch installation (1.28 mg/kg vs. 1.27 mg/kg; *p* = 0.9752) (Fig. 3).

**Figure 3.**
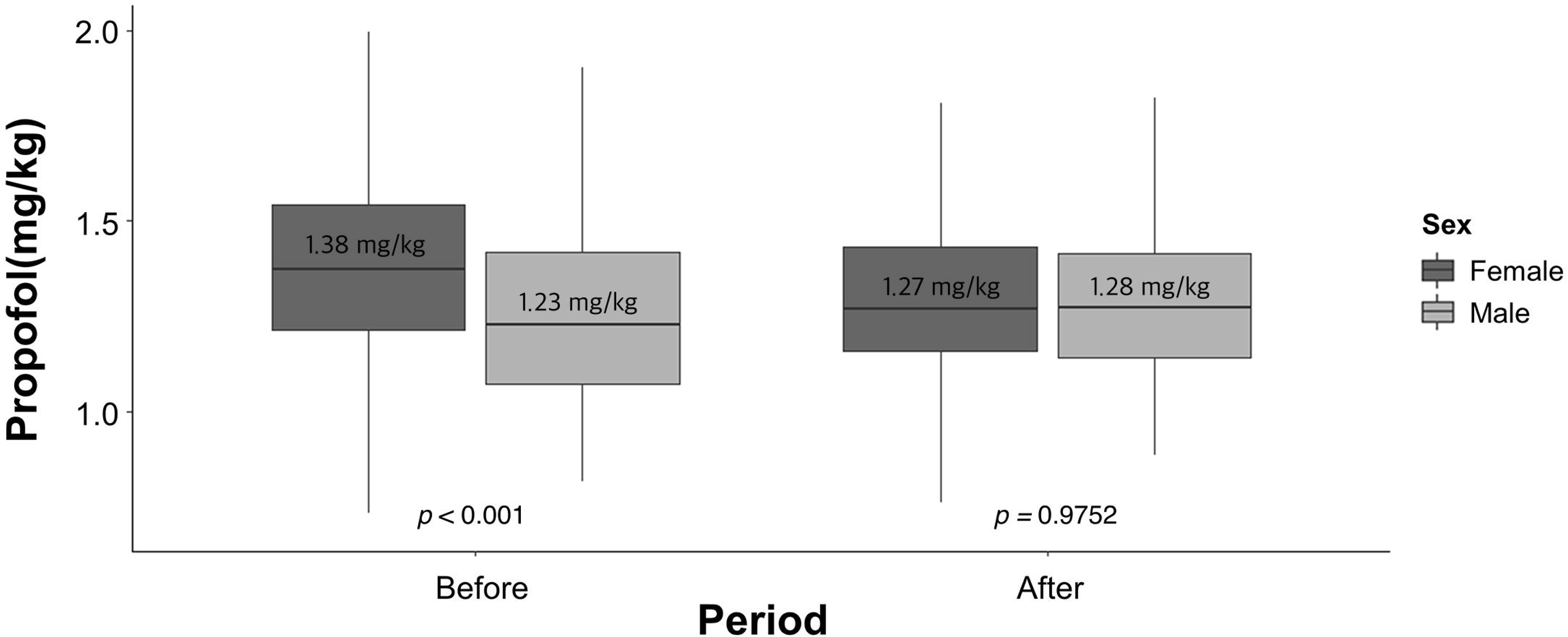
Box plot of the PW for both sexes in the before and after group.

## Discussion

Propofol is known as a safe drug, and it has been used for sedation endoscopy since the mid-1980s and has been used by many endoscopists since the 1990s[25–27]. However, if propofol is used in high doses for a long time, serious adverse events, such as propofol infusion syndrome, may occur[28]. Even with a slight overdose, pulmonary and cardiovascular side effects, such as suppression of spontaneous respiration or lowering of cardiac output, can occur easily. Of note, propofol overdose currently has no antidote.

Due to the large variance in the required dose of propofol among individuals, it is necessary to start with the smallest dosage and gradually reach the desired level of sedation through additional administration. In this case, the dosing interval is very important. If the dosing interval is too short, an overdose could occur[19]. On the contrary, if the dosing interval is too long, the patient’s sedation level will decrease, resulting in arousal. Therefore, in order to make the dosing interval appropriate, the endoscopist must be aware of the passage of time accurately.

When a person become anxious, he or she cannot correctly perceive the passage of time and thought that time is passing quickly[20,29]. This event is also true for upper gastrointestinal endoscopists who simultaneously manage the patient’s vital signs and depth of sedation during an endoscopic procedure, requiring a higher level of concentration throughout the entire procedure. Generally, a conventional watch enables us to perceive the passage of time. However, when faced with high cognitive loads, relying on multi-step methods, like subtracting the previous time from the current time, to recognize the passage of time becomes more prone to errors. Consequently, additional drug administration is more likely to proceed earlier than normal, indicating a too narrow interval, thereby possibly leading to overdose. The stopwatch can help intuitively in perceiving the passage of time without such calculations and make drug injection easy at proper intervals. Hence, using a stopwatch allows a more appropriate amount of the drug to be administered at the right time.

### Limitations

Generally, the arm-brain circulation time is known to be around 15-20 seconds, and propofol begins to act on the brain after approximately 20 seconds from administration. Therefore, both men and women initiated the examination or attempted additional drug administration after 20-30 seconds post propofol injection. However, the arm-brain circulation time could slightly differ between men and women. Looking at the results of this study, the increase in propofol usage in men could be due to the longer arm-brain circulation time in men, which might have led to an earlier decision for additional administration. It would be a good approach to assume different arm-brain circulation times for men and women in future prospective studies for comparison.

Despite the implementation of the stopwatch, the results of our study revealed a significant decrease in propofol requirement only among females, while an increase in propofol requirement was observed in males. This outcome may be seen as a limitation of our study since it did not demonstrate a consistent decrease in propofol requirement across both genders. However, despite the increase in propofol usage among males, it can be noted that the propofol usage per body weight became similar between males and females after the introduction of the stopwatch, suggesting that a meticulously maintained drug interval can potentially lead to more uniform and appropriate propofol dosing across diverse patient groups. This observation is supported by the histograms (Fig. 4A, Fig. 4B), which indicate a shift towards more normalized patterns in males after the introduction of the stopwatch, contrasting with the previously discrete patterns seen in males compared to females.

**Figure 4.**
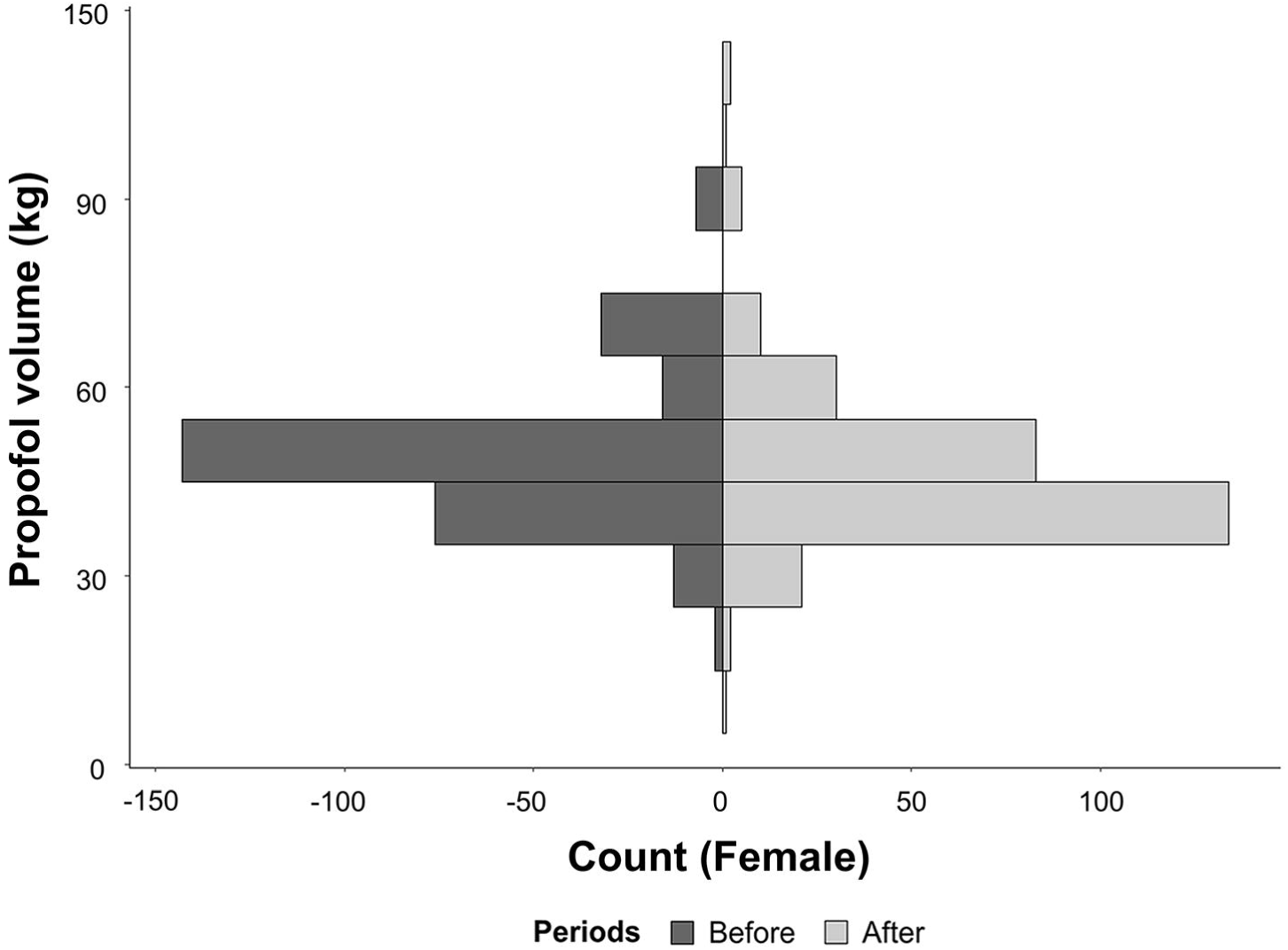
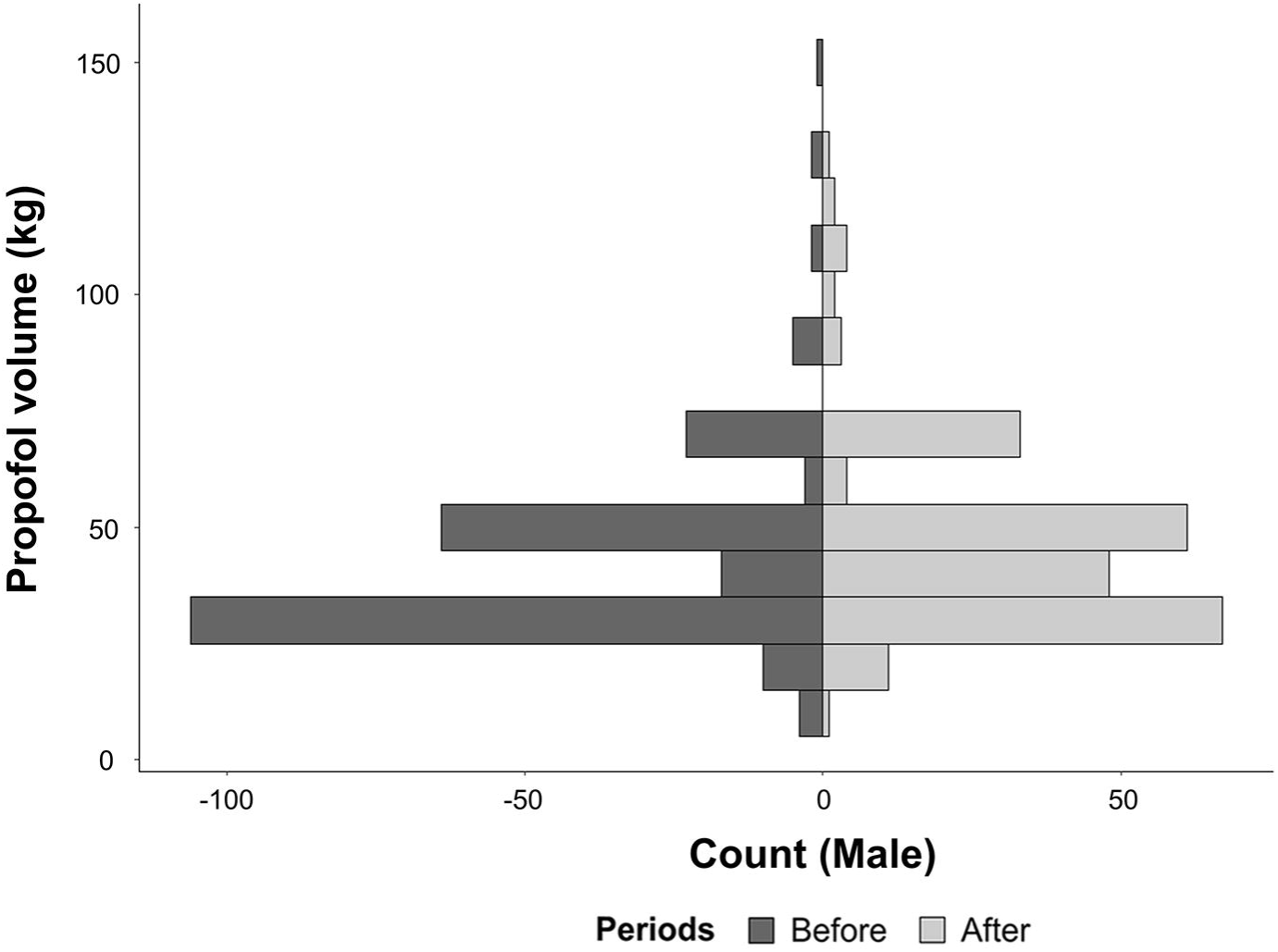
A, Histogram of propofol used in before and after group in females. B, Histogram of propofol used in before and after group in males.

Furthermore, these findings, which indicate no difference in propofol usage between males and females, contradict the consistent findings from previous studies that suggested higher propofol requirements in females [22,23,30–32]. However, it is important to note that most studies highlighting higher propofol requirements in females were primarily focused on general anesthesia, and not on short-term sedation as investigated in this study. Rather, Horiuchi et al. found no significant difference in the dose of propofol between males and females when a small amount of propofol was used in their study.[13]. Therefore, it is possible that the propofol dosing requirement (PW) does not exhibit significant differences between males and females, particularly in the context of upper gastrointestinal endoscopy procedures with shorter durations.

The fact that this study was conducted by a single endoscopy specialist contributes to maintaining homogeneity in the research. However, it is believed that this could have a negative impact on generalizing the research findings. And also, this study is retrospective in nature, and thus there may be potential insufficiencies in controlling variables across groups. Therefore, it is hoped that prospective studies involving multiple endoscopy specialists from various centers will be conducted. This will allow for obtaining more comprehensive and representative results.

Despite these limitations, this study is meaningful in that simple and basic actions have led to significant changes in drug use. When endoscopists alone must perform both sedation endoscopy and patient monitoring simultaneously, a stopwatch may be helpful in reducing the cognitive burden of endoscopists. Although the results of these positive changes may not be apparent in both sexes, the endoscopist’s work stress is expected to reduce, indicating a potential positive effect on patient safety.

### Conclusions

This study illustrates that the implementation of a simple tool such as a stopwatch during sedation endoscopy can significantly influence propofol dosage patterns, leading to a decrease in propofol usage in females and an increase in males, yet resulting in more uniform dosing per body weight across genders, suggesting a potential for improved precision in administration and enhancing patient safety during the procedure.

## Data Availability

All relevant data are within the manuscript and its Supporting Information files.

## Acknowledgments

The author would like to thank Enago (www.enago.co.kr) for the English language review.

